# Deep longitudinal phenotyping of wearable sensor data reveals independent markers of longevity, stress, and resilience

**DOI:** 10.1101/2020.12.24.20248672

**Authors:** Timothy V. Pyrkov, Ilya S. Sokolov, Peter O. Fedichev

**Affiliations:** Gero PTE, 60 Paya Lebar Road # 05-40B Paya Lebar Square 409051 Singapore; Moscow Institute of Physics and Technology, 141700, Institutskii per. 9, Dolgoprudny, Moscow Region, Russia

**Keywords:** Biological Age Acceleration (BAA), Public Health, Personalized Interventions, Gompertz Law, Aging, Resilience

## Abstract

Biological age acceleration (BAA) models based on blood tests or DNA methylation emerge as a *de facto* standard for quantitative characterizations of the aging process. We demonstrate that deep neural networks trained to predict morbidity risk from wearable sensor data can provide a high-quality and cheap alternative for BAA determination. The GeroSense BAA model presented here was tolerant of gaps in the data, and exhibited a superior association with life-expectancy over the average number of steps per day, e.g., in groups stratified by professional occupations. The association between the BAA and effects of lifestyles, the prevalence or future incidence of diseases was comparable to that of BAA from models based on blood test results. Wearable sensors let sampling of BAA fluctuations at time scales corresponding to days and weeks and revealed the divergence of organism state recovery time (resilience) as a function of chronological age. The number of individuals suffering from the lack of resilience increased exponentially with age at a rate compatible with Gompertz mortality law. We speculate that due to stochastic character of BAA fluctuations, its mean and auto-correlation properties together comprise the minimum set of biomarkers of aging in humans.

## INTRODUCTION

Any advances in personalized and informed lifestyle interventions to promote longevity and health will require a reliable and immediate feedback on health status changes in response to treatments. Such capabilities have just recently became available in the form of biological clocks and are increasingly used in the field of quantitative aging research. State-of-art implementations involve machine learning of the associations of the DNA methylation patterns [1] or blood variables [2–4] with either the chronological age or risks of death and diseases. The aging clocks have been used in clinical trials of anti-aging interventions [5].

Large scale biochemical or genomic profiling of Biological age acceleration (BAA) is, however, still logistically difficult and expensive. Mobile technology holds a great promise for democratization of population health studies. It already provides engagement tools to help customers maintain physical activity levels, body weight, and adhere to lifestyles known to promote a healthy lifespan. In 2019, one-in-five U.S. adults (21%) report they regularly use a wearable fitness tracker or smartwatch [6]. The health and home fitness app downloads grew by 46% during COVID-19 lockdown [7].

In fact, only the mobile technology can provide adequate sampling rates for truly longitudinal large-scale epidemiological studies. Recent examples include the analysis of the worldwide distribution of physical activity [8], changes in physical activity levels in response to COVID-19 lockdown [9], and the associations of physical activity and the risks of COVID-19 mortality [10, 11]. There are, however, multiple unresolved issues, such as inaccuracies of sensor data, missing data, outliers, varying measurements between devices of different manufacturers, and seasonal variation of physical activity [12, 13] - all precluding from wider acceptance of the wearables signal in population studies.

We applied deep learning technology to systematically address these challenges. We trained and characterized a simple model that learns physical activity patterns from wearable devices, which are directly associated with morbidity risks on the population level. Accordingly, the organism state representation output by this model is a single dynamic variable closely related to BAA. The neural network architecture included components specifically designed to resolve the missing data and solve transferability across platforms. We found that both blood-based and wristband step-counter-based models demonstrated surprisingly similar levels of sensitivity in applications involving BAA associations with diseases and lifestyles. Moreover, the activity-based models’ signal-to-noise ratio could be improved by averaging over longer motion tracks. After just a few months of averaging, the activity-based model applied to a wristband signal may detect the effects of chronic diseases and smoking at the same level of significance as blood-based PhenoAge from [2] and Dynamic Organism State Indicator (DOSI) from [4]. Same finding held for the association of BAA with the incidence and severity of seasonal infectious diseases (including COVID-19).

Finally, we investigated the auto-correlation properties of the BAA fluctuations. The diverging autocorrelation times are typical for systems approaching tipping or disintegration point [14] and a hallmark of aging [4, 15]. Accordingly, we observed vanishing recovery rate and the exponentially increasing fraction of individuals with long recovery times in subsequent age cohorts. The number of non-resilient individuals doubled every 8 years, which is compatible with the mortality rate doubling time characteristic to the Gompertz mortality law [16]. We conclude that due to the inherent stochastic character of BAA fluctuations, the BAA mean and the BAA autocorrelation time (the resilience) are two the most basic and independent health indicators, closely related to aging and human mortality.

## RESULTS

### Biological age predicts morbidity and mortality

We trained the GeroSense system, a deep artificial neural network (Fig. 1) to extract health-associated features from the physical activity recordings. The system included the encoder part, which took the input in the form of a series of step count per minute measurements for at least as long as one week and compressed the signal into 4-dimensional representations (embeddings). During the training procedure, these vectors were further fed into the domain-adaptation network, trained to reduce the difference between the feature sets distribution in samples originating from different devices. In such a way, we were able to produce the most common features present in the motion data.

**Figure 1.**
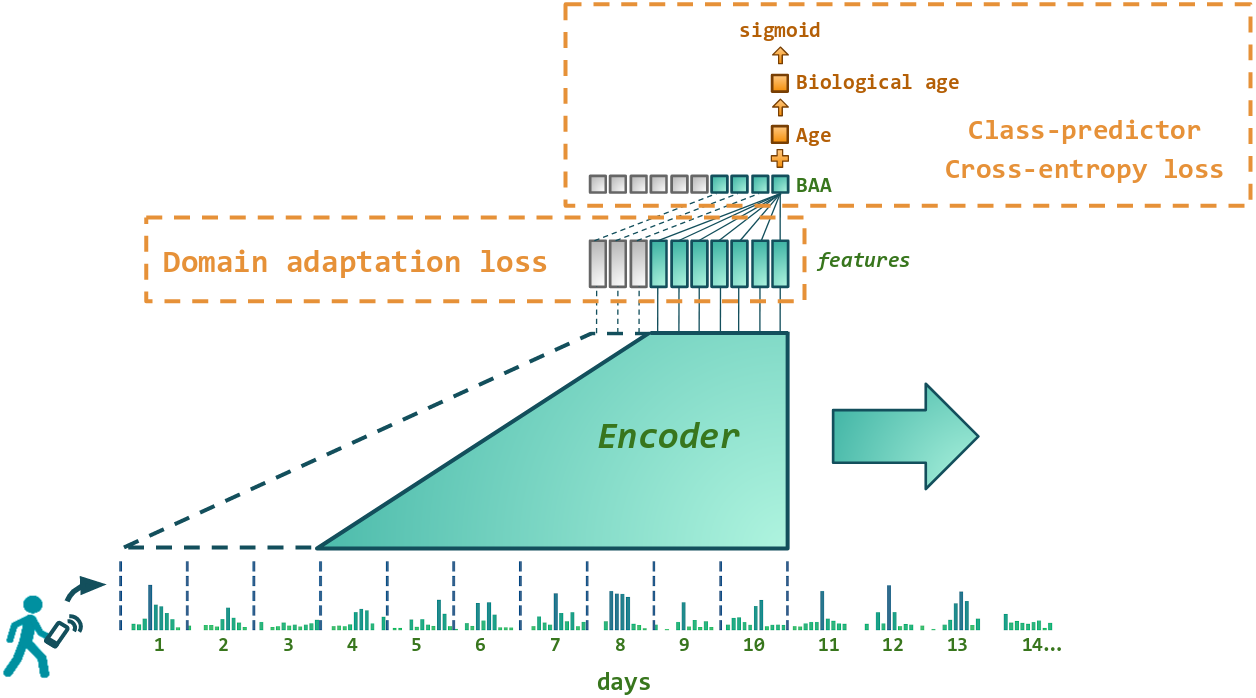
Architecture of the neural network predicting biological age acceleration (BAA). GeroSense model predicts BAA once per day based on step counts recorded by wearable or mobile device sensors using each individual’s week-long physical activity tracks. The network components responsible for the feature extraction and BAA output are shown in green. BAA can be predicted for any sample of arbitrary length exceeding one week. For example, BAA on the day 10 is predicted using the step counts data coming from the day 4 through the day 10, and so forth. Shown in red are the network components used only during the training procedure. One is the discriminator responsible for domain adaptation between e.g. smartphones and smartwatches. The other is the class predictor based on the log-odds ratio trained to predict morbidity binary status for UK Biobank and NHANES.

At the top layer, log-linear proportional hazards models of all-cause mortality are natural tools to build the biological age acceleration models, see, e.g., PhenoAge model [2, 17]. If, however, the number of observed events is small, a simple logistic regression model provides an excellent approximation to the solution of the corresponding proportional hazards [18, 19]. Therefore, in the present study, we trained the neural network using cross-entropy loss to predict binary labels: the prevalence of at least one chronic disease. Overall, we labeled events for 23% and 29% samples in NHANES and UKB, respectively (see Materials and Methods section “Morbidity status” for the precise definition).

The model’s output was the Biological Age Acceleration (BAA), output once per each seven days and calculated as the linear combination of the physical activity signal embeddings and biological sex label. During the training procedure, BAA was added to the chronological age of each participant to produce biological age followed by sigmoid activation layer and cross-entropy loss on prediction of morbidity status.

To control for over-fitting, we split all data into training and test subsets. The quality of GeroSense BAA for predicting the morbidity status was similar in training and test subsets in both NHANES (Fig. 2A) and UK Biobank (Fig. 2C) with ROC AUC 0.60 − 0.61 in test subsets.

**Figure 2.**
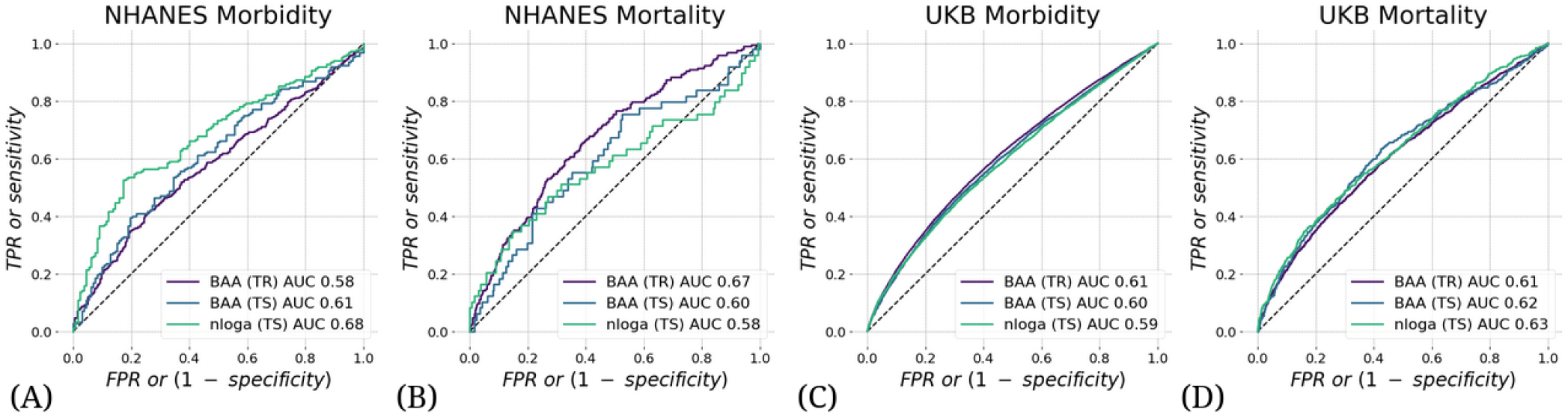
Biological age acceleration (BAA) scores mortality and morbidity events. BAA estimated from patterns of intraday changes in physical activity level is associated with the morbidity and mortality in NHANES (**A, B**) and UK Biobank (**C, D**) datasets. The performance was tested in participants aged 45 − 75 y.o. and was similar in training and test subsets.

We also expected the high concordance between the mortality and morbidity predictors [20]. Accordingly, we tested the ability of the model to predict future mortality events (see Figs. 2B and 2D for the summary of the GeroSense BAA model performance in NHANES and UKB datasets, respectively). The scoring performance was similar to that of morbidity status and yielded ROC AUC 0.60 − 0.62 in test subsets.

### BAA and the life expectancy in professional occupation groups

BAA from the network was superior to average daily physical activity-based BAA in scoring life expectancy in various professional occupations. The number of steps per day averaged over a sufficiently long period is an easy-to-understand and adjustable parameter that predicts mortality and morbidity [20]. This can be readily seen in Fig. 2, where the negative logarithm of the number of daily steps (nloga) has all the properties required of BAA. However, the average physical activity obviously cannot be a good biological age measure. It is strongly affected by social factors and working schedule and therefore has poor correlation with life expectancy across countries [8] and between groups of different professional occupation (Fig. 3A).

**Figure 3.**
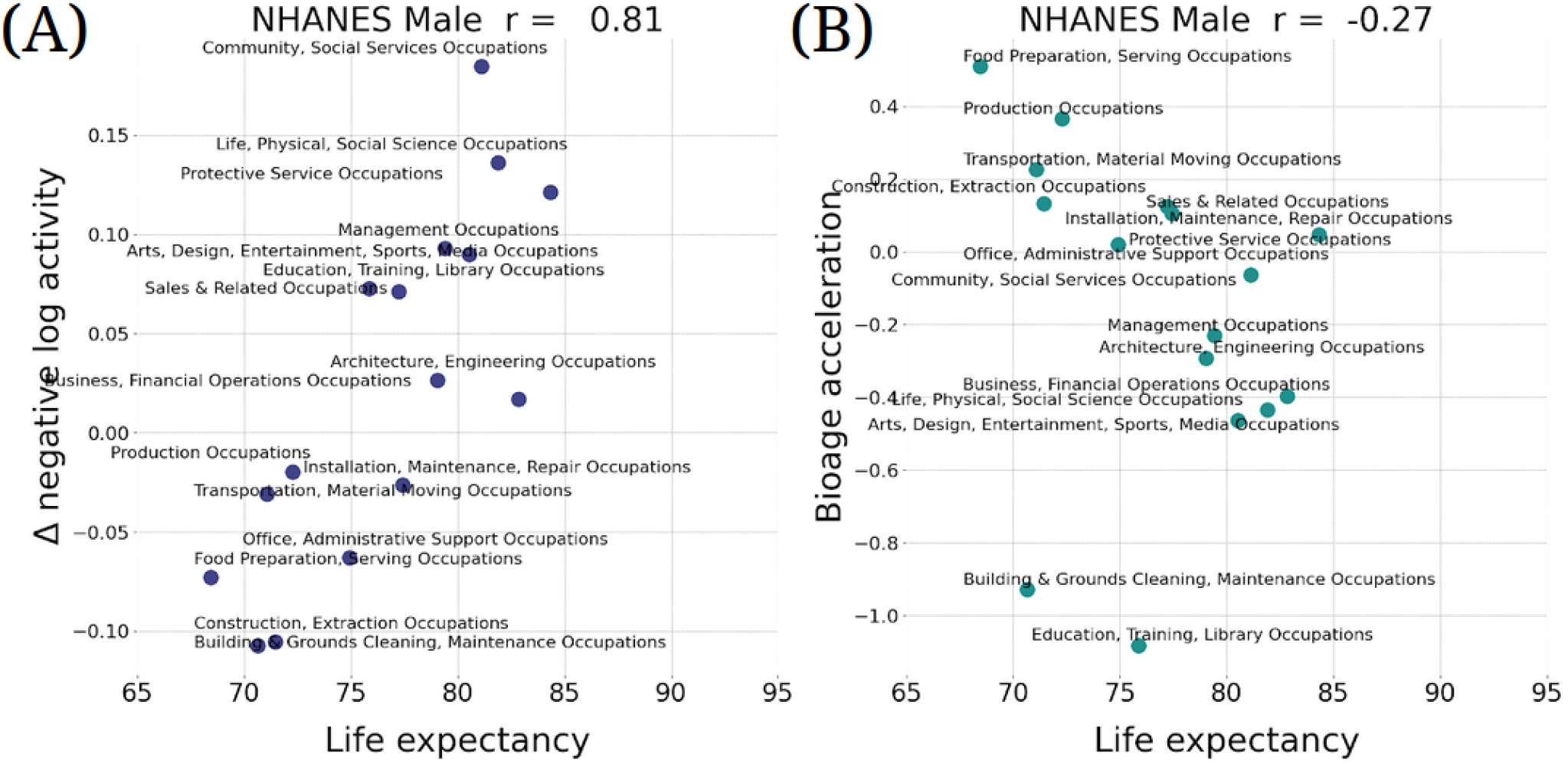
Biological age acceleration (BAA) correctly ranks life expectancy. **A**) Assuming negative logarithm of average daily steps is a proxy for bioage, the observed positive correlation (Pearson’s *r* = 0.81 for males) with life expectancy is an incorrect prediction. **B**) The negative correlation (Pearson’s *r* = −0.27 fro males) of GeroSense BAA wth life expectancy is correct. Similar results were observed for females with Pearson’s *r* = 0.19 and *r* = −0.55, respectively (data not shown). The calculations were performed in NHANES 2005 − 2006 cohorts aged 30 − 60 y.o.

Notably, the GeroSense system produced BAA from wearable sensors data, which properly ranked professional occupation groups in NHANES according to both genders’ empirical life expectancy. We did not have access to and hence could not test the association of physical activity and lifespan data across countries. Therefore, Gerosense BAA’s ability to score the life-expectancy of populations of different countries remains an open issue.

### Cross-platform transferability of BAA and seasonal variations

The embeddings of physical activity tracks depend on the signal source, whether it is a smartphone or a smartwatch. Deep Neural networks are powerful feature-extraction tools and a proper choice to address this issue. We employed the domain adaptation network minimizing the feature-wise Kullback–Leibler divergence loss between samples originating from different devices during the training procedure. The problem is akin to batch removal. The proposed procedure helped the GeroSense network to learn the features most common between UKB, NHANES, and samples obtained from iPhone and Apple Watch.

Seasonal changes affect blood parameters [21], and physical activity patterns recorded by wearables [12]. The seasonal variations of the activity patterns may be an additional source of unwarranted fluctuations of the biological age estimates. We applied another Kullback–Leibler divergence minimization to penalize pair-wise differences in distributions of features for UK Biobank samples collected in the summer and winter.

The domain adaptation worked well: BAA level distributions were almost indistinguishable between the samples originating from smartphones and smartwatches (*p* = 2*E* − 5). In contrast, the levels of negative logarithm of average physical activity were much more different (*p* = 2.7*E* − 80). The difference was expected but is still striking since we analyzed the smartphone and smartwatch data from the same users.

The results of the statistical testing (p-values) strongly depend on the sample size. That is why, here and in all the following examples, we report p-values obtained for the same maximum size of 500 in each group. The p-values themselves are calculated using Fisher’s combined probability test (see details in Materials and Methods section).

Notably, there was a very significant drop in the physical activity levels during the COVID-19 pandemic lockdown in March through May 2020 as compared to the same period in 2019 (*p* < 1*E* 3 − 0 for nloga). This was consistent with what was reported earlier [9]. In contrast, the increase in BAA was much less significant (*p* > 1*E* 10). This may indicate that BAA responds weaker to the lockdown than the expected decrease in physical activity.

The decreased average level of physical activity (nloga) was associated with the increased COVID-19 risk in UKB [11], although it was not clear if this was not an effect of chronic diseases burden (also known for its association with increased BAA). In Fig. 6 we report that the excess BAA predicted the increased risk of COVID-19 (HR = 2.4, *p* = 4*E* − 2 for 18 of UKB subjects died from the disease) in the subset of randomly sampled 500 UK Biobank participants free of chronic diseases at the time of measurements (2013 − 2015).

### Side-by-side comparison of motion data- and blood-based aging clocks

We compared performance of different BAA models for stratification of cohorts of NHANES participants of various health status and lifestyles. We have already seen in physical activity data [22] that the disease and smoking labels are associated with elevated BAA among individuals without chronic diseases. In our tests, the sensitivity of the BAA derived from blood markers was comparable to that of the self-reported questionnaire. GeroSense BAA performed consistently well in the same set of tests and conditions, see Figs. 4 and 5.

**Figure 4.**
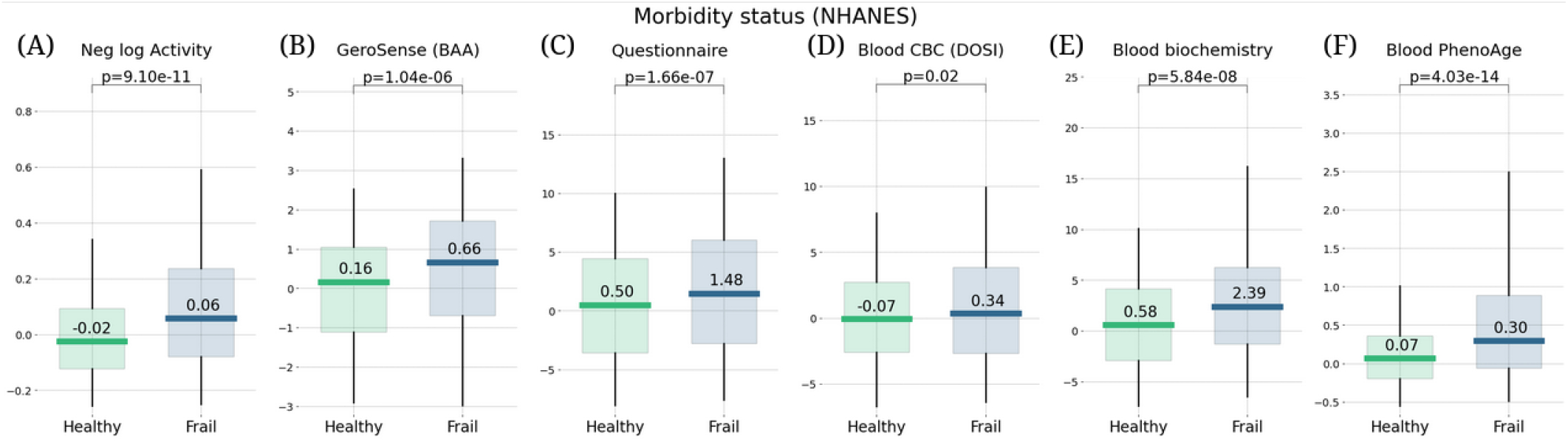
Morbidity status scored by wearable BAA and blood-based bioage models. BAA and chronic diseases: **A**) the negative logarithm of daily step counts, **B**) GeroSense BAA **C**) questionnaire [22], **D**) CBC-based DOSI [4], **E**) log-mortality risk trained using combined CBC and Blood biochemistry variables, and **F**) Blood-based PhenoAge [2]. The plots are produced for NAHNES users aged 45 − 75 y.o.

**Figure 5.**
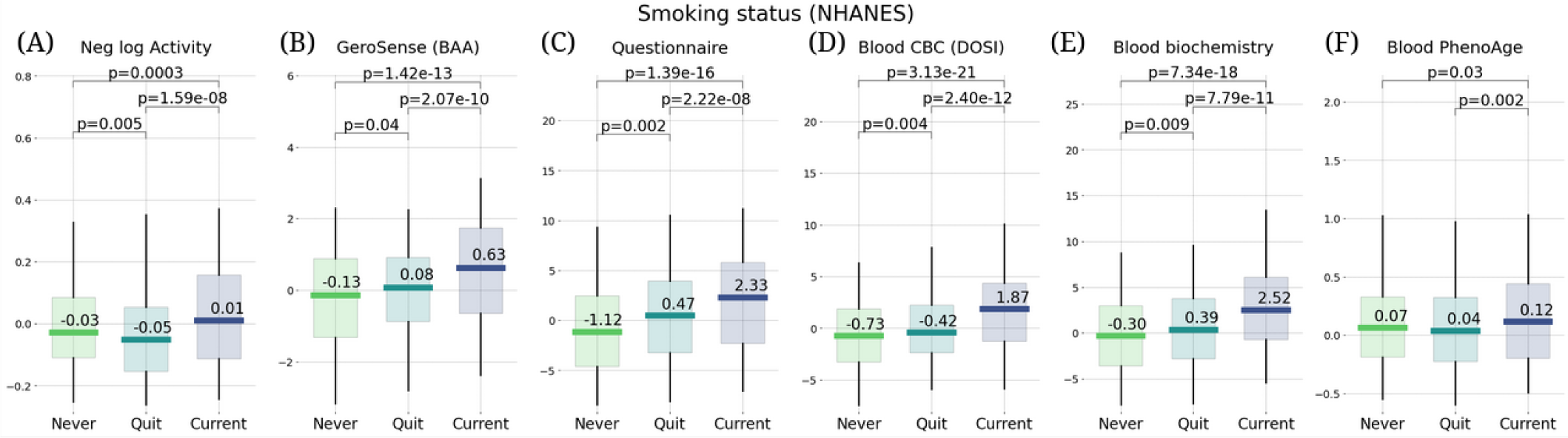
Smoking status representing an unhealthy lifestyle scored by wearable BAA and blood-based bioage models. BAA and smoking: **A**) BAA in the form of the negative logarithm of daily step counts, **B**) GeroSense BAA, **C**) questionnaire [22], **D**) CBC-based DOSI [4], **E**) log-mortality risk trained using combined CBC and Blood biochemistry variables **F**) PhenoAge [2]. The plots are produced for NAHNES users aged 45 − 75 y.o.

**Figure 6.**
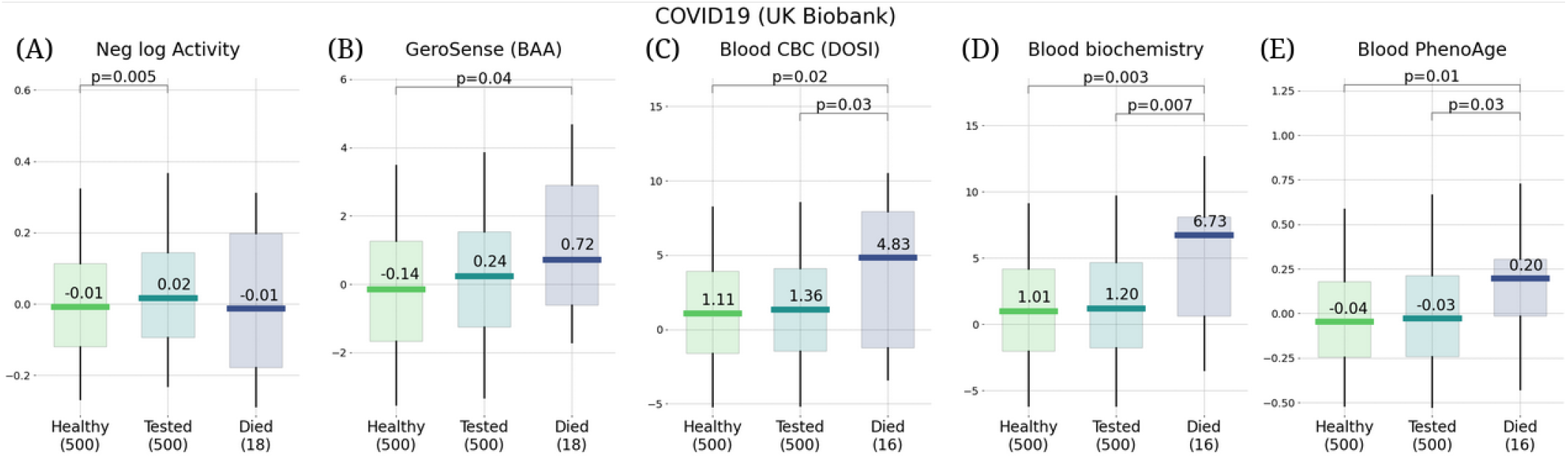
Future risks of COVID-19 scored by wearable BAA and blood-based bioage models. Association of BAA with the future incidence of COVID-19: **A**) BAA in the form of the negative logarithm of daily step counts, **B**) GeroSense BAA, **C**) CBC-based DOSI [4], **D**) CBC and Blood biochemistry hazards model, and **E**) Blood-based PhenoAge [2]. All data are given for UK Biobank users aged 45 − 75 y.o.

Estimation of the BAA from wearable sensors has an advantage over blood-based models. It arises from its ability to further improve the signal to noise ratio by averaging over sufficiently long motion data streams. We demonstrated this with self-reported morbidity and smoking status provided by smartphone app and wrist-band tracker users.

Averaging of GeroSense BAA predictions over a few-weeks long tracks led to a dramatic improvement of association between the BAA and morbidity/smoking status (Fig. 7A). As expected, the sensitivity of the model was comparatively lower once we used smartphones instead of wristbands as the source of the data (Fig. 7B) but also improved upon averaging over several weeks.

**Figure 7.**
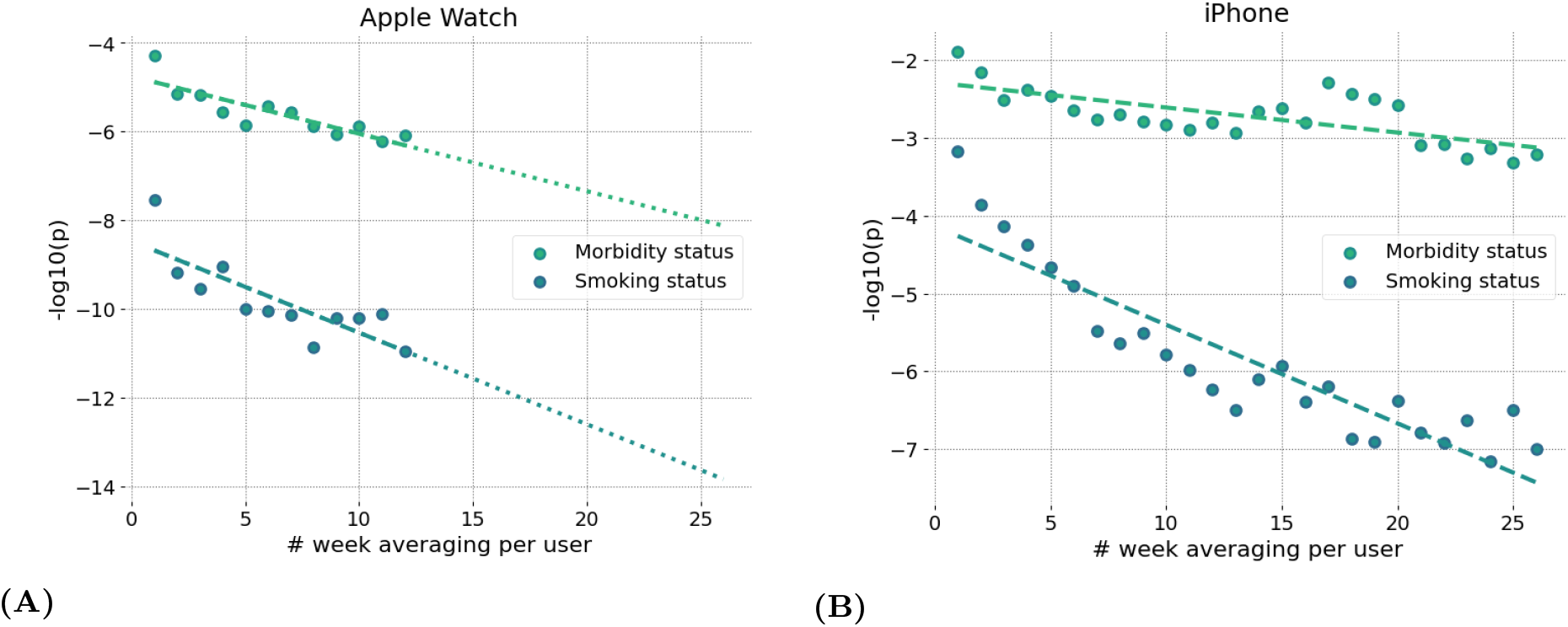
Accuracy of BAA grows with longer data collection intervals. Significance of association of GeroSense BAA with morbidity and current smoking status is improved when BAA is averaged over several weeks of data obtained from sensors of smartwatch (**A**) and smartphone (**B**).

### Longitudinal analysis of BAA fluctuations reveal age-dependent loss of resilience

BAA reversibly depends on lifestyles. Hence, BAA is a dynamic variable more characteristic of stress rather than aging and responding to random organism state perturbations in a stochastic manner. We used longitudinal tracks of step counts from Fitbit devices and calculated the autocorrelation function for every user. The autocorrelation function decayed exponentially. Accordingly, we carried out the exponential fit to infer the autocorrelation time as a measure of recovery rate or resilience. This quantity is a natural quantitative measure of an organism’s ability to recover its equilibrium state after stress.

The characteristic decay time was typically in the range of a few weeks and increased with age. Fig. 8A shows the dependence of the recovery rate (the inverse auto-correlation time) on chronological age. The graph was produced by averaging over age-stratified cohorts and resembles much what we have previously reported for blood-based marker DOSI [4]. The recovery rate decreased approximately linearly with age, indicating the effective loss of resilience at some age exceeding 100 y.o. The same extrapolation would suggest that the recovery time increases approximately hyperbolically and would diverge at the same age, indicating the complete loss of resilience and the dynamic stability of the organism state.

**Figure 8.**
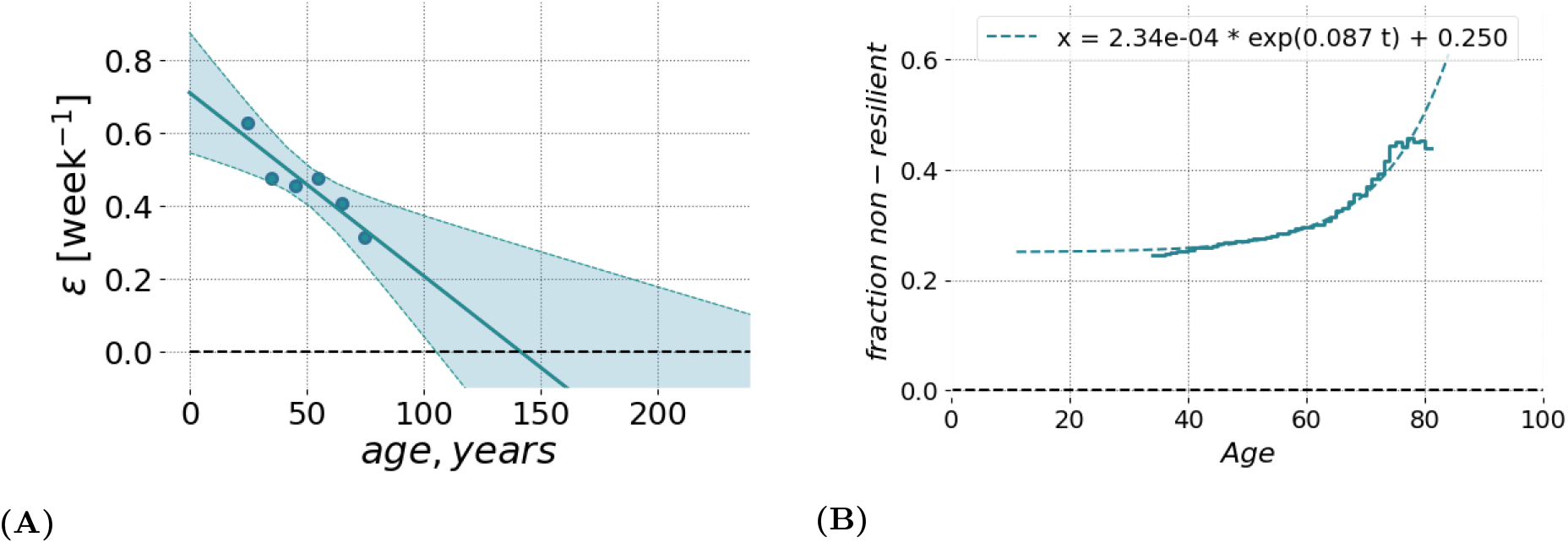
Resilience and its age-related degradation can be measured using longitudinal motion sensor data. **A**) The relaxation rate (or the inverse characteristic recovery time) computed for sequential age-matched cohorts of Fitbit users decreased approximately linearly with age. The recovery rate could be extrapolated to zero in the age exceeding ∼ 110 y.o. (at this point, we may expect the complete loss of resilience and, hence, loss of stability of the organism state). The shaded area shows the 95% confidence interval of fit using GeroSense BAA. **B**) The fraction of individuals suffering from the lack of resilience (defined as BAA’s autocorrelation time exceeding 3 weeks; the vertical axis) as the function of chronological age (the horizontal axis). The autocorrelation time was computed from longitudinal tracks of GeroSense BAA predicted for Fitbit wristband users.

To further investigate the relationship between resilience and aging, we identified individuals, which failed to recover quickly under stress. We established a somewhat arbitrary resilience cutoff corresponding to the recovery time exceeding 3 weeks. The fraction of such “non-resilient” individuals increased exponentially as a function of age (see Fig. 8B). Moreover, this grwoth demonstrated the characteristic doubling rate of 0.087 per year, which was close to the mortality rate doubling rate according to the Gompertz mortality law.

## DISCUSSION

We report the development and characterization of a deep neural network model trained to quantify the state of human health from the analysis of intraday physical activity tracks collected by consumer wearable devices (including mobile phones). The quantity has properties of biological age acceleration (BAA): it is associated with chronic diseases and life-shortening lifestyles, predicts the risks of death and future incidence of chronic diseases in cohorts of individuals free of chronic diseases [4].

Deep neural networks are natural tools for learning non-trivial and highly non-linear representations of the input data. Convolutional and recurrent networks have been used for the analysis of intraday physical activity data streams from wearable devices and predictive modeling of health outcomes [23] including biological age [17, 24]. Often such models demonstrate a moderate improvement in accuracy at a price of a decreased transferability across datasets with different baseline feature levels. This is, of course, is well-known batch effect problem in large scale studies in biology [25], which is often aggravated by feature-rich deep learning architectures [13, 26].

GeroSense BAA model employs additional neural network components to address this domain shift problem to insure learning device-independent representations of input signal. To achieve this goal, we imposed an additional loss in the course of training to penalize model parameters if distributions of learned representations were too far apart for data from different domains (devices). Without such a domain adaptation, the properties of the signal may indeed be very different even in the same biological context. For example, the (log-scaled) average number of daily steps recorded by phone was significantly lower (*p* = 2.7*E* − 80) than that by the smartwatch in the data from the same users. GeroSense BAA network successfully resolved this batch effect and yielded essentially indistinguishable BAA distributions for the same population (*p* = 2*E* − 5).

The average activity level recorded by the same device in a group of people of the same gender, professional occupation, and country of residence is already an excellent and a popular proxy to biological age. The association between the mean activity and health is robust and hence is the basis for the popular recommendation to take a minimum of 10, 000 steps a day [27]. However, the average activity level is highly context-dependent, which is why it is poorly associated with life expectancy across countries [8]. In our study, we demonstrate that the average activity is incorrectly (negatively) associated with the life expectancy across professional occupation groups (Fig. 3A).

The device-independent features from intraday physical activity patterns from the GeroSense network are still associated with health but decoupled from the mean activity. The procedure did not undermine the predictive power of GeroSense model, as we could see from the BAA association with mortality events (Fig. 2). GeroSense BAA was superior in scoring life expectancy in professional occupation subgroups (Fig. 3B). This feature of the model should be useful in applications involving health risk assessment and life insurance applications.

Biological clocks based on mortality risk, including GeroSense BAA, are associated with the prevalence of chronic diseases (Fig. 4) and life-shortening lifestyles, such as smoking, in a reversible way (Fig. 5). This is totally consistent with earlier observations of the effect of smoking on physical activity [20], blood markers [4, 22], and DNAm PhenoAge [2].

In NHANES cohorts, the GeroSense model produced the association between the BAA and the morbidity and smoking labels at the significance level matching that of the BAA calculated based on self-reported health questionnaire [22], blood test-based bioage including CBC only [4], and blood biochemistry [22], and Phenotypic Age [2].

The longitudinal character of motion data provides a natural way to improve the signal-to-noise ratio by averaging over sufficiently long tracks (see Figs. 7A and 7B). This may be critical for mobile phone applications since the step counts recorded by phones suffer from missing data whenever a device is idle and is not recording the user’s walks. Our analysis suggests that GeroSense BAA from smartphones can be averaged to a useful level once at least few months of data are available for an individual. The inferior performance of the biological age model in smartphone data can be mitigated by smartphone population coverage compared to that of wristband wearable devices. The smartphone motion data can be used for truly large-scale epidemiological studies involving cohort comparisons. The latter factor might turn important to mitigate the issues of non-representative datasets due to possible income/health status [13, 28] and already observed enrollment biases [29].

We observed, that GeroSense BAA is also associated with the incidence of non-chronic diseases. This is consistent with earlier observations of the association of lower physical activity levels and risks of COVID-19 infection [10, 11], although it was not clear whether this is an effect of chronic diseases, also negatively affecting mobility. GeroSense BAA was better associated with the incidence of COVID-19 than the average physical activity level in UKB among a sub-population of individuals free of chronic health conditions (Fig. 6).

The average physical activity dropped worldwide in 2020 in the course of COVID-19 lockdown [9]. We also observed a significant change in (log-scaled) number of daily step counts in our data, but not in GeroSense BAA during March - May 2020 as compared to the same period in 2019. We provided evidence suggesting that GeroSense BAA more efficiently sores those at risk of getting an infection than the physical activity level. The effects of lockdown on morbidity risk may be smaller, than one could expect simply monitoring the drop of the activity. Further studies including direct association with epidemiological data are required to test this hypothesis.

The idea of reducing complex biological signals to as little as one variable, the BAA, in relation to the current or future health arises from the effectively low dimensionality of physiological systems. Typically, physiological and behavioral responses manifest themselves as highly coordinated changes in physiological variables, such as blood tests [4] or daily physical activity patterns [20]. The increasing concordance between the physiological indices is expected to increase late in life, as the range of the fluctuations and the organism state recovery time effectively diverge at advanced ages indicating a maximum attainable lifespan [4]. On the contrary, the number of the relevant variables is expected to increase if we turn to characterization of the organism state variation at a higher sampling rate. This might be the case for situation involving response to an acute illness on time scales of days or a few weeks [30], such as increased RHR during fever [31, 32] or change in sleep patterns as potentially a COVID-19 specific signal [33, 34].

The quantitative characterization of the dynamic properties of BAA fluctuations or recovery processes requires a reliable determination of baseline BAA. This task may be hampered by seasonal variation of the physiological state variables, such as blood tests [21, 35], blood pressure [36], resting heart rate [37], and of course physical activity [12]. High-quality research acknowledge this problem and adjust for baseline oscillations [28]. Such corrections are straightforward for relatively short time scales involved in acute respiratory illnesses [30] or post-operative recovery [38].

Unfortunately, proper adjustments are not always possible in practice. Health outcomes associated with BAA may be years apart from the time (and hence the season) of observations [19]. Otherwise, the time of measurements may be available at poor granularity. For example, NHANES provides publicly only the binary labels corresponding to the winter-spring (November-April) or summer-fall (May-October) seasons.

We trained the GeroSense BAA model with an additional loss penalizing the winter-summer distribution difference. In such a way, the model output is decoupled from seasonal variations and yet demonstrated pretty good performance in ranking health outcomes. We expect that this feature of GeroSense BAA will be handy for practical applications.

The longitudinal character of motion data allows the investigation of organism state fluctuations in response to natural stresses and diseases. We computed autocorrelation functions of GeroSense BAA along the individual BAA trajectories. Recovery rate measured as the inverse decay time of the autocorrelation function demonstrated age-dependent decrease (Fig. 8A). Extrapolation to advanced ages shows, that the recovery rate vanishes (and hence the resilience formally diverges) at some age exceeding 100 years, which may be an indication of limiting lifespan [4].

The recovery time in the most healthy individuals was in the range of a few weeks. We used a somewhat arbitrary cutoff corresponding to the recover rate less than 3 week^_^−1^_^ and used it to mark individuals with longer recovery time as those who lack resilience. We observed a progressive exponential increase of the fraction of non-resilient persons in the population with age (Fig. 8B). This number grew and doubled every 8 years, which is close to the mortality rate doubling time from Gompertz mortality law for human population [16].

Long auto-correlation times of state fluctuations are typical for complex systems approaching a tipping point or in the process of disintegration [14] and is a hallmark of aging [4, 15]. Case fatality rates (CFR) accelerate with age in case of COVID-19, stroke, and probably other diseases. The characteristic doubling rate in the case of COVID-19 is reported in [39] as 6 − 9 years. Our estimation from the figure in [40] yielded ≈ 10 years for the doubling rate for one-year survival of stroke patients. The physiology of the infection and stroke diseases is apparently very different. The similar patterns of age-dependence of CFR is intriguing and may suggest that the loss of resilience may be a good marker of the approaching loss of dynamic stability of an organism and hence a major and universal contributing factor to the fatality.

The reversible character of the association between mortality-risk based BAA and unhealthy lifestyles (such as smoking) suggests that BAA is not a biomarker of aging but instead is a measure of the overall stress level. BAA’s dependence on age in large cross-sectional datasets is a marker of stress imposed by the increasing burden of chronic diseases. The high sampling rate achievable by the motion data lend us a richer set of biomarkers associated with age. Aside from the average BAA level, the continuous data collected by wearable sensors provides a practical opportunity to investigate the autocorrelation and variance properties of BAA fluctuations, which are independent organism state variables, each uniquely informing about the user’s health state. We can hardly imagine large scale blood test study involving sampling more often than once a month or so for healthy people. Therefore, only the motion data analysis exemplified here is the only technology currently up for the task.

Wearable device motion data have already been used for monitoring acute illnesses including detection of early signs of the outbreak of influenza-like illnesses [28] and COVID-19 [30, 34]. Application of motion data, including wider deployment of the GeroSense system, described here, should provide means to monitor levels of stress and resilience in response to environmental conditions or interventions on a population level. We hope that future developments will lead to further applications of AI in geroscience research, public health and policy decision-making.

## Data Availability

The data that support the findings of this study is available upon reasonable request from the authors.

## ACKNOWLEDGEMENTS

This research has been conducted using data from UK Biobank, a major biomedical database (UK Biobank website: www.ukbiobank.ac.uk; UK Biobank project ID 21988).

## MATERIALS AND METHODS

### Datasets

#### UK Biobank

Physical activity for UK Biobank participants aged 40 − 80 y.o. (54777 female and 42543 male) was measured by Axivity AX3 tri-axial accelerometers worn on the wrist for one week. We converted 100Hz raw acceleration measurements to step counts per minute to fit the format of data in other datasets used in this study. Number of steps during each consecutive minute was counted as the number of peaks of the absolute value of acceleration exceeding 1.3*g*. To ensure the local noise does not affect the result, only one peak (the highest) was counted in each 480*ms* sliding window with a step of 160*ms* resulting in at most 3 step counts during each 960*ms*. Steps closer to each other than 90*s* were combined into walking bouts and bouts with less than 5 steps in total were discarded.

#### NHANES

Physical activity for NHANES 2005 − 2006 participants was used in the form of step counts per minute collected by ActiGraph AM-7164 single-axis accelerometer worn on hip. Data were retrieved from the file “Physical Activity Monitor” of Examination data category. Samples for 3362 female and 3148 male participants aged 6 − 85 y.o. were used.

#### Healthkit

Physical activity for users of Gero app aged 45 − 75 y.o. (464 female and 1412 male users of smartphone, 125 female and 598 male users of smart-watch) was obtained from Healthkit. Raw activity data comprised number of steps recorded by either smart-phone or smartwatch during a time period with start and end timestamps and was resampled to equispaced time series of steps per minute.

### Morbidity status

Binary morbidity status for Healthkit dataset was assigned according to response to survey question “Have you ever been told you have one of the following: diabetes, hypertension, cancer, coronary heart disease, heart failure, heart attack, or stroke?” Binary morbidity status of NHANES and UK Biobank participants was assigned according to presence of at least one of those diagnoses. We used NHANES data on health condition and age at diagnosis available in the questionnaire category “Medical Conditions” (MCQ). Data on diabetes and hypertension was retrieved additionally from questionnaire categories “Diabetes” (DIQ) and “Blood Pressure & Cholesterol” (BPQ), respectively. For UK Biobank we aggregated ICD10 (block level) data to match that of NHANES and used the following ICD10 codes to cover the health conditions in UK Biobank: diabetes (E10-E14), hypertension (I10-I15), cancer (C00-C99), coronary heart disease (I20-I25), congestive heart failure (I50), myocardial infarction (I21, I22), and stroke (I60-I64).

### Life expectancy

Empirical life expectancy from birth was determined for professional occupation groups using linked death register follow-up data for NHANES 2005 − 2015 surveys. To do that we fitted parameters of Gompertz likelihood adopted from [41]:

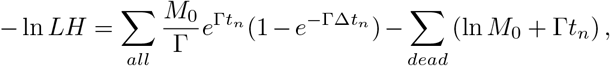

where *M*_0_ and Γ are the initial mortality rate and mortality doubling rate of the Gompertz mortality law, *t*_*n*_ is the age of *n*-th participant at the end of followup, and Δ*t*_*n*_ is the follow-up time since enrollment in NHANES survey.

Once *M*_0_ and Γ were obtained by the fit for each professional occupation group, the life expectancy 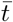 in the group was calculated as:

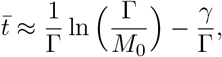

where *γ* ≈ 0.58 is the Euler–Mascheroni constant. The expression for life expectancy is asymptotically correct whenever *M*_0_*/*Γ ≪ 1, which is definitely true in human cohorts.

### Statistical analysis

Statistical analysis of association of various Biological Age measures with morbidity/smoking status was performed using two-sided Mann-Whitney test. To ensure the reported p-values are comparable between tests we used the same cutoff of maximum 500 samples in each test with 100 random samplings followed by combining p-value according to Fisher’s combined probability method [42]. All statistical tests were carried out using python package SciPy (version 1.5.2).

### Neural network Architecture

Deep neural network architecture is schematically shown in Figure 1. Wearable data is input in the form of continuous array of steps per minute. The input is immediately converted to a one-hot embedding representation, where each bin corresponds to an increment of 4 steps per minute. Next, the encoded data is processed by a block of 16 1D-convolutional layers, each having 16 filters with kernel size of 3 and “elu”-activation. One in two convolutional layer is followed by a local max-pooling with stride 2, 3 or 5, and each layer followed by batch-normalization. The output of the convolutional block was 4 features per each 1440 points in the input array, which corresponds to the number of minutes in one day. Finally, the features were subject to a 7 day long average pooling and linearly combined with binary biological sex label so that the deep neural network was capable of outputting a prediction once per day based on 7 previous days.

The output of the deep neural network was interpreted as the Biological Age Acceleration (BAA) expressed in years of healthy life expectancy gained or lost. To guarantee this, during the supervised training of class label predictor we obtained the value of Biological age of each NHANES and UK Biobank user by adding the network output (BAA) to the chronological age. The Biological age was then subject to sigmoid activation and fitted to binary morbidity status label, assuming that such procedure is an approximation to fitting proportional hazards model [18, 19].

The Domain adaptation networks were employed in the from of pairwise Kullback-Leibler divergence loss functions applied to enforce similar feature distributions for samples from UK Biobank on one side and NHANES, HealthKit smartphones and smartwatches on the other side. Additionally a domain adaptation was applied to UK Biobank samples collected during summer and winter.

The training procedure was run for 2000 iterations, each batch comprising 256 samples. Class predictor was trained on each iteration for UK Biobank samples and only on one in five iterations for NHANES to avoid potential overfitting since the number of NHANES samples was small. All domain adaptation networks were trained on each iteration. Each network was trained using Adam optimizer as implemented in python package tensorflow-gpu (version 2.3.1) with learning rate of 1*E* − 3.

## References

[1] Steve Horvath. Dna methylation age of human tissues and cell types. Genome biology, 14(10):3156, 2013.

[2] Morgan E Levine, Ake T Lu, Austin Quach, Brian H Chen, Themistocles L Assimes, Stefania Bandinelli, Li-fang Hou, Andrea A Baccarelli, James D Stewart, Yun Li, et al. An epigenetic biomarker of aging for lifespan and healthspan. Aging (Albany NY), 10(4):573, 2018.

[3] Evgeny Putin, Polina Mamoshina, Alexander Aliper, Mikhail Korzinkin, Alexey Moskalev, Alexey Kolosov, Alexander Ostrovskiy, Charles Cantor, Jan Vijg, and Alex Zhavoronkov. Deep biomarkers of human aging: Application of deep neural networks to biomarker development. Aging (Albany NY), 8(5):1021–33, May 2016.

[4] Timothy V Pyrkov, Konstantin Avchaciov, Andrei E Tarkhov, Leonid I Menshikov, Andrei V Gudkov, and Peter O Fedichev. Longitudinal analysis of blood markers reveals progressive loss of resilience and predicts ultimate limit of human lifespan. bioRxiv, page 618876, 2019.

[5] Gregory M Fahy, Robert T Brooke, James P Watson, Zinaida Good, Shreyas S Vasanawala, Holden Maecker, Michael D Leipold, David TS Lin, Michael S Kobor, and Steve Horvath. Reversal of epigenetic aging and immunosenescent trends in humans. Aging cell, 18(6):e13028, 2019.

[6] EA Vogels. About one-in-five americans use a smart watch or fitness tracker. Washington, DC: Pew Research Centre, 2019.

[7] Carmen Ang. The growth of home fitness apps. Visual Capitalist, 2020.

[8] Tim Althoff, Jennifer L Hicks, Abby C King, Scott L Delp, Jure Leskovec, et al. Large-scale physical activity data reveal worldwide activity inequality. Nature, 547(7663):336, 2017.

[9] Geoffrey H Tison, Robert Avram, Peter Kuhar, Sean Abreau, Greg M Marcus, Mark J Pletcher, and Jeffrey E Olgin. Worldwide effect of covid-19 on physical activity: a descriptive study. Annals of internal medicine, 2020.

[10] Xiaomeng Zhang, Xue Li, Ziwen Sun, Yazhou He, Wei Xu, Harry Campbell, Malcolm G Dunlop, Maria Timofeeva, and Evropi Theodoratou. Physical activity, bmi and covid-19: an observational and mendelian randomisation study. medRxiv, 2020.

[11] Kejun Ying, Ranran Zhai, Timothy V Pyrkov, Marco Mariotti, Peter O Fedichev, Xia Shen, and Vadim N Gladyshev. Genetic and phenotypic evidence for the causal relationship between aging and covid-19. medRxiv, 2020.

[12] Tessa Strain, Katrien Wijndaele, Paddy C Dempsey, Stephen J Sharp, Matthew Pearce, Justin Jeon, Tim Lindsay, Nick Wareham, and Søren Brage. Wearable-device-measured physical activity and future health risk. Nature Medicine, 26(9):1385–1391, 2020.

[13] Jennifer L Hicks, Tim Althoff, Peter Kuhar, Bojan Bostjancic, Abby C King, Jure Leskovec, Scott L Delp, et al. Best practices for analyzing large-scale health data from wearables and smartphone apps. NPJ digital medicine, 2(1):1–12, 2019.

[14] Marten Scheffer, Jordi Bascompte, William A Brock, Victor Brovkin, Stephen R Carpenter, Vasilis Dakos, Hermann Held, Egbert H Van Nes, Max Rietkerk, and George Sugihara. Early-warning signals for critical transitions. Nature, 461(7260):53–59, 2009.

[15] D Podolskiy, I Molodtcov, A Zenin, V Kogan, LI Menshikov, Vadim Gladyshev, Robert J Shmookler Reis, and PO Fedichev. Critical dynamics of gene networks is a mechanism behind ageing and gompertz law. arXiv preprint arXiv:1502.04307, 2015.

[16] Benjamin Gompertz. On the nature of the function expressive of the law of human mortality, and on a new mode of determining the value of life contingencies. Philosophical transactions of the Royal Society of London, 115:513–583, 1825.

[17] Timothy V Pyrkov, Konstantin Slipensky, Mikhail Barg, Alexey Kondrashin, Boris Zhurov, Alexander Zenin, Mikhail Pyatnitskiy, Leonid Menshikov, Sergei Markov, and Peter O Fedichev. Extracting biological age from biomedical data via deep learning: too much of a good thing? Scientific reports, 8(1):5210, 2018.

[18] Manfred S Green and Michael J Symons. A comparison of the logistic risk function and the proportional hazards model in prospective epidemiologic studies. Journal of chronic diseases, 36(10):715–723, 1983.

[19] Robert D Abbott. Logistic regression in survival analysis. American Journal of Epidemiology, 121(3):465–471, 1985.

[20] Timothy V. Pyrkov, Evgeny Getmantsev, Boris Zhurov, Konstantin Avchaciov, Mikhail Pyatnitskiy, Leonid Menshikov, Kristina Khodova, Andrei V. Gudkov, and Peter O. Fedichev. Quantitative characterization of biological age and frailty based on locomotor activity records. Aging, 10(10):2973–2990, 2018.

[21] Bian Liu and Emanuela Taioli. Seasonal variations of complete blood count and inflammatory biomarkers in the us population-analysis of nhanes data. PloS one, 10(11):e0142382, 2015.

[22] Timothy V Pyrkov and Peter O Fedichev. Biological age is a universal marker of aging, stress, and frailty. In Biomarkers of Human Aging, pages 23–36. Springer, 2019.

[23] Tom Quisel, David C Kale, and Luca Foschini. Intra-day activity better predicts chronic conditions. arXiv preprint arXiv:1612.01200, 2016.

[24] Syed Ashiqur Rahman and Donald A Adjeroh. Deep learning using convolutional lstm estimates biological age from physical activity. Scientific reports, 9(1):1–15, 2019.

[25] Jeffrey T Leek, Robert B Scharpf, Héctor Corrada Bravo, David Simcha, Benjamin Langmead, W Evan Johnson, Donald Geman, Keith Baggerly, and Rafael A Irizarry. Tackling the widespread and critical impact of batch effects in high-throughput data. Nature Reviews Genetics, 11(10):733–739, 2010.

[26] Alan A Cohen, Vincent Morissette-Thomas, Luigi Ferrucci, and Linda P Fried. Deep biomarkers of aging are population-dependent. Aging (Albany NY), 8(9):2253, 2016.

[27] Bernard CK Choi, Anita WP Pak, and Jerome CL Choi. Daily step goal of 10,000 steps: a literature review. Clinical & Investigative Medicine, 30(3):146–151, 2007.

[28] Jennifer M Radin, Nathan E Wineinger, Eric J Topol, and Steven R Steinhubl. Harnessing wearable device data to improve state-level real-time surveillance of influenza-like illness in the usa: a population-based study. The Lancet Digital Health, 2(2):e85–e93, 2020.

[29] A. Ganna and E. Ingelsson. 5 year mortality predictors in 498,103 UK Biobank participants: a prospective population-based study. Lancet, 386(9993):533–540, Aug 2015.

[30] Aravind Natarajan, Hao-Wei Su, and Conor Heneghan. Assessment of physiological signs associated with covid-19 measured using wearable devices. medRxiv, 2020.

[31] Jouko Karjalainen and Matti Viitasalo. Fever and cardiac rhythm. Archives of internal medicine, 146(6):1169–1171, 1986.

[32] Xiao Li, Jessilyn Dunn, Denis Salins, Gao Zhou, Wenyu Zhou, Sophia Miryam Schüssler-Fiorenza Rose, Dalia Perelman, Elizabeth Colbert, Ryan Runge, Shannon Rego, et al. Digital health: tracking physiomes and activity using wearable biosensors reveals useful health-related information. PLoS biology, 15(1):e2001402, 2017.

[33] Tejaswini Mishra, Meng Wang, Ahmed A Metwally, Gireesh K Bogu, Andrew W Brooks, Amir Bahmani, Arash Alavi, Alessandra Celli, Emily Higgs, Orit Dagan-Rosenfeld, et al. Pre-symptomatic detection of covid-19 from smartwatch data. Nature Biomedical Engineering, pages 1–13, 2020.

[34] Giorgio Quer, Jennifer M Radin, Matteo Gadaleta, Katie Baca-Motes, Lauren Ariniello, Edward Ramos, Vik Kheterpal, Eric J Topol, and Steven R Steinhubl. Wearable sensor data and self-reported symptoms for covid-19 detection. Nature Medicine, pages 1–5, 2020.

[35] M Reza Sailani, Ahmed A Metwally, Wenyu Zhou, Sophia Miryam Schüssler-Fiorenza Rose, Sara Ahadi, Kevin Contrepois, Tejaswini Mishra, Martin Jinye Zhang, Lukasz Kidzinski, Theodore J Chu, et al. Deep longitudinal multiomics profiling reveals two biological seasonal patterns in california. Nature communications, 11(1):1–12, 2020.

[36] Kwang-il Kim, Nima Nikzad, Giorgio Quer, Nathan E Wineinger, Matthieu Vegreville, Alexis Normand, Nicolas Schmidt, Eric J Topol, and Steven Steinhubl. Real world home blood pressure variability in over 56,000 individuals with nearly 17 million measurements. American journal of hypertension, 31(5):566–573, 2018.

[37] Giorgio Quer, Pishoy Gouda, Michael Galarnyk, Eric J Topol, and Steven R Steinhubl. Inter-and intraindividual variability in daily resting heart rate and its associations with age, sex, sleep, bmi, and time of year: Retrospective, longitudinal cohort study of 92,457 adults. Plos one, 15(2):e0227709, 2020.

[38] Ernesto Ramirez, Nikki Marinsek, Benjamin Bradshaw, Robert Kanard, and Luca Foschini. Continuous digital assessment for weight loss surgery patients. Digital Biomarkers, 4(1):13–20, 2020.

[39] Didac Santesmasses, José Pedro Castro, Aleksandr A. Zenin, Anastasia V. Shindyapina, Maxim V. Gerashchenko, Bohan Zhang, Csaba Kerepesi, Sun Hee Yim, Peter O. Fedichev, and Vadim N. Gladyshev. Covid-19 is an emergent disease of aging. Aging Cell, 19(10):e13230, 2020.

[40] Tom Skyhoj Olsen, Zorana Jovanovic Andersen, and Klaus Kaae Andersen. Age trajectories of stroke case fatality: leveling off at the highest ages. Epidemiology, pages 432–436, 2011.

[41] Ralf Bender, Thomas Augustin, and Maria Blettner. Generating survival times to simulate cox proportional hazards models. Statistics in medicine, 24(11):1713–1723, 2005.

[42] Ronald Aylmer Fisher et al. Statistical methods for research workers. Statistical methods for research workers., (5th Ed), 1934.

